# Joint associations of parental personality traits and socioeconomic position with trajectories of offspring depression: findings from up to 6925 families in a UK birth cohort

**DOI:** 10.1101/2020.08.19.20177899

**Authors:** Tim Cadman, Alex S. F. Kwong, Paul Moran, Heather O’Mahen, Iryna Culpin, Deborah A Lawlor, Rebecca M. Pearson

## Abstract

**Background:** Parental personality may influence the course of offspring depression but this is unclear. It is also unknown whether the impact of parental personality on offspring depression is moderated by socioeconomic position (SEP). Our aims were to describe trajectories of depressive symptoms across adolescence for offspring of parents with and without maladaptive personality traits and to test for effect modification by SEP.

**Methods:** A longitudinal study in the Avon Longitudinal Study of Parents and Children birth cohort (ALSPAC; ns = 3054 to 7046). Exposures were binary measures of maladaptive parental personality traits and the outcome was depressive symptoms ages 11 to 24 (SMFQ; range 0 – 26).

**Results:** Offspring of mothers with high maladaptive traits showed higher levels of depressive symptoms at all ages (SMFQ difference at age 10 = 0.66, CI 0.25 – 1.28, p = 0.02; age 22 = 1.00, CI 0.51 – 1.50, p < 0.001). There was weaker evidence of an association between paternal maladaptive personality traits and offspring depressive symptoms (SMFQ difference at age 10 = 0.21, CI –0.58 – 0.99, p = 0.60; age 22 = 0.02, CI –0.94, 0.90, p = 0.97). We found no consistent evidence of effect modification by SEP.

**Conclusions:** Offspring of mothers with high levels of maladaptive personality traits show evidence of greater depressive symptoms throughout adolescence although the absolute increase in symptoms is small. Evidence for the effect of fathers’ personality was weaker. Socio-economic position and maladaptive personality traits appear to be independent risk factors.

## Introduction

There is a large body of evidence showing that parental mental health problems are associated with an increase in the risk of a range offspring mental health problems. (Stein et al., 2014) The majority of the research has focused on outcomes associated with maternal depression, especially in the perinatal period. (Goodman et al., 2011) However, emerging evidence suggests that other parental traits such as maladaptive personality traits may also represent important risk factors for offspring mental health (Pearson et al., 2018)

Maladaptive personality traits include domains such as emotional dysregulation, suspicion, hostility and impulsivity. Whilst continuously distributed, (Hopwood et al., 2018) at the extreme end these traits cluster together in the clinical syndromes described as borderline and antisocial personality disorders. In contrast to depressive symptoms, (which often show a pattern of remission and possible relapse) these traits are relatively stable and inflexible self and behavioural characteristics and may affect offspring mental health development in the absence of other mental health problems. (Eyden, Winsper, Wolke, Broome, & MacCallum, 2016) Findings from cross-sectional studies of parents with Borderline Personality Disorder (BPD) have reported elevated levels of depressive symptoms in adolescent offspring. (Abela, Skitch, Auerbach, & Adams, 2005; Barnow, Spitzer, Grabe, Kessler, & Freyberger, 2006) Recently, in a well-powered longitudinal study, we reported the existence of associations between maladaptive maternal personality traits and elevated levels of depression, anxiety and self-harm in their offspring at age 18. (Pearson et al., 2018)

The mechanisms leading to elevated depressed symptoms in offspring of parents with maladaptive personality traits are likely to include both genetic and environmental factors. Heritability of BPD estimated at 40% (Amad, Ramoz, Thomas, Jardri, & Gorwood, 2014) and there is evidence that depression and maladaptive personality traits share common genetic correlates. (Witt et al., 2017) In terms of environmental mechanisms, a key area of focus has been on parenting and the early attachment relationship. Parenting is a highly demanding and stressful task, requiring perspective taking and the regulation of one’s own emotions in the service of the child’s well-being. Parents with maladaptive personality traits may become more quickly emotionally dysregulated and be more likely to respond with less optimal strategies. (Eyden et al., 2016) Indeed, a number of cross-sectional studies have reported parenting differences in this population, including higher levels of parental hostility, (Herr, Hammen, & Brennan, 2008) intrusiveness, (Hobson et al., 2009) lower sensitivity, (Crandell, Patrick, & Hobson, 2003; Newman, Stevenson, Bergman, & Boyce, 2007) and over-protectiveness among mothers with BPD. (Elliot et al., 2014; Feldman et al., 1995) One longitudinal study also found that self-report of over-protective and rejecting parenting styles partially mediated the relationship between mother and offspring BPD symptoms, though the size of the effect was modest. (Reinelt et al., 2014)

### Limitations of previous research

Most studies examining the mental health of offspring of parents with high levels of these personality traits are cross-sectional, (Eyden et al., 2016) and the few longitudinal studies have assessed offspring outcomes at only one time point. (Pearson et al., 2018) However, it is important to understand how parental personality impacts the manifestation of depressed mood *over time*, to inform when and how children may be most at risk. For example, it is important to understand whether such traits influence overall levels of depression at any time, or alternatively whether they are associated with steeper rises in depressed mood during sensitive periods in the life course, such as during adolescence. (Crone & Dahl, 2012) Understanding the nature of this relationship could be important for developing critically-timed interventions for individuals with parents with personality difficulties.

A further limitation is that few studies have explored the joint associations of socioeconomic factors and parent personality traits with offspring mental health. Understanding the contextual factors associated with the risk of offspring mental health problems in the presence of parental personality traits is important from a public health perspective. (Mikkonen, Moustgaard, Remes, & Martikainen, 2016) Furthermore, there is evidence that socioeconomic position may moderate associations of parental mental health with offspring mental health. For example, in our previous work, maternal education appeared to moderate the association of maternal post-natal depressive symptoms with offspring depression but not that of antenatal depression. (Pearson et al., 2013) Similarly, a meta-analysis, which included 65,619 mother-offspring pairs from 121 studies, reported that the relationship between maternal depression (at any point after birth) and offspring internalising in childhood was strongest in lower socioeconomic samples. (Goodman et al., 2011) However there was significant heterogeneity in effect sizes between studies in the high & low SEP groups, and it was not reported how consistently interaction effects were found within studies. Contrary to these findings, a large (N = 138,559 mother-offspring pairs) cohort study published since this meta-analysis found maternal depression (experienced between child ages 9 – 14) and low SEP to be independent risk factors for later offspring depression (measured ages 15–20) with no statistical evidence of an interaction. (Mikkonen et al., 2016) The lack of consistent evidence for socioeconomic position modifying the association of maternal depression with offspring depression may be due to different time periods when maternal depression is measured, or because the small number of studies finding evidence of interaction are chance findings.

Finally, few studies have examined the association of paternal maladaptive personality traits on child outcomes. Paternal associations with offspring outcomes are important because (with the exception of sex-linked effects) fathers and mothers contribute equally to genetic risk, yet may differ in caregiving and roles. (McKinney & Renk, 2007) Evidence on the association of paternal perinatal depression with offspring depression is inconsistent. Whilst some studies have observed positive associations of paternal depression with offspring mental health, (Ramchandani, Stein, Evans, & O’Connor, 2005) (Kvalevaag et al. 2013, others find no evidence of an association or reported weaker associations in comparison to maternal depression. (Pearson et al., 2013) (Stein et al., 2014). We previously found a close to null association of maladaptive paternal personality with offspring anxiety, depression and self-harm measured once when the offspring were 18 years old (Pearson et al., 2018); herein we extend this research by exploring the associations of maternal and paternal maladaptive personality traits on offspring depression symptoms trajectories from childhood to early adulthood. The aims of this study therefore were:

1. To describe separately trajectories of depressive symptoms of offspring of (i) mothers and (ii) fathers with high levels of maladaptive personality traits, and compare these to trajectories of offspring with parents without these traits
2. To explore whether any association between parental maladaptive personality traits and offspring depressive symptoms is moderated by SEP.

## Method

### Sample

The sample comprised participants from the Avon Longitudinal Study of Parents and Children (ALSPAC), an ongoing population-based study. The study website contains details of all data available through a fully searchable data dictionary and variable search tool http://www.bristol.ac.uk/alspac/researchers/our-data/). Ethical approval for the study was obtained from the ALSPAC Ethics and Law Committee and the Local Research Ethics Committees. Informed consent for the use of data collected via questionnaires and clinics was obtained from participants following the recommendations of the ALSPAC Ethics and Law Committee at the time Pregnant women resident in Avon, UK with expected dates of delivery 1st April 1991 to 31st December 1992 were invited to take part in the study. The initial number of pregnancies enrolled is 14,541 (for these at least one questionnaire has been returned or a “Children in Focus” clinic had been attended by 19/07/99). Of these initial pregnancies, there was a total of 14,676 foetuses, resulting in 14,062 live births and 13,988 children who were alive at 1 year of age. (For further details on the cohort profile, representativeness, and phases of recruitment, see (Boyd et al., 2013; Fraser et al., 2013; Northstone et al., 2019).

### Measures

#### Exposure

##### Parental maladaptive personality traits

Maladaptive personality traits in mothers and fathers were assessed using the Karolinska Scales of Personality (KSP) inventory (Gustavsson, 1997) when their child was mean age 9 years. The KSP is a self-report questionnaire containing 135 questions measuring 15 personality traits relevant to psychological functioning and vulnerability to psychiatric conditions. The majority of the subscales relate to neuroticism; however as neuroticism is strongly linked to depression and we wanted to capture a distinct construct, we focused on 5 subscales identified *a priori* (Monotony Avoidance, Impulsivity, Verbal Anger, Suspicion and Detachment, number of items = 47) which may be theoretically distinct from the neuroticism domain and which are reflective of relational and affect dysregulation. Based on our previous work showing that the accumulation of high levels of multiple traits is associated with greater offspring mental health risk, (Pearson et al., 2018) parents were categorised as having high maladaptive personality if they scored in the top quartile for at least 3 out of the 5 selected scales.

#### Outcomes

##### Depressive symptoms: Short mood and feelings questionnaires (SMFQ)

Depressive symptoms were assessed using the Short Mood and Feelings Questionnaire (SMFQ) (Angold, Costello, Messer, & Pickles, 1995) on up to 9 occasions between the ages of 11 and 24 years. The SMFQ is a 13-item questionnaire that measures the occurrence of depressive symptoms over the preceding two weeks with higher scores indicating more severe depressive symptoms (range 0–26). Questionnaires were completed by the offspring either via postal/internet questionnaires or by computer at a clinic visit.

#### Potential effect modifiers

##### Socio-economic position (SEP)

Two indicators were chosen to capture different aspects of SEP: (i) education and (ii) material hardship.

Parental education is strongly related to future income and employment and also reflects non-material family resources (e.g knowledge) (Galobardes, Shaw, Lawlor, Lynch, & Davey Smith, 2006). Binary variables for maternal and paternal education were created from parents’ report of their highest educational qualification at the time of the mothers’ recruitment in early pregnancy. High education was defined as Advanced Level (A-level) or comparable qualifications (A-levels are exams taken in up to 4 subjects usually at age 18 years and required for university entry and some skilled jobs) or University degree. Low education was defined as no qualifications or qualifications below A-level.

Material hardship was assessed within the first year of the child’s life for both parents using the question: “How difficult at the moment do you find it to afford these items for the child: food, clothing, heating, rent, items for child”. Possible responses ranged from 0 (“not difficult”) to 3 (“very”), giving a total range of 0–15 across the five items. Binary variables were created for each parent created using a cutoff of ≥ 5 corresponding to material hardship scores in the top 20% of the sample. (Joinson, Kounali, & Lewis, 2017). This aspect of SEP has previously been shown in ALSPAC to be associated with offspring depression. (Joinson et al., 2017)

#### Confounders

Confounders were selected *a priori* on the basis that they were known to, or plausibly, influence parental maladaptive personality traits and offspring depression: parental age at child birth (years), maternal drinking in period of pregnancy (yes/no), parental depression during the postnatal period taken as the average score on the Edinburgh Postnatal Depression Scale (EPDS) measured at 2 months and 8 months postpartum as used in previous studies, (Stein et al., 2014), smoking in pregnancy/at the time of partners pregnancy (yes/no) and self-report of experiencing intimate partner violence (yes/no). We also adjusted for offspring sex as it is associated with depression and this adjustment may have improved the precision of our results.

### Data analysis

#### Trajectories of depressive symptoms for offspring of mothers and fathers with maladaptive personality traits

Trajectories of depressive symptoms were estimated using multilevel growth-curve modelling. (Bryk & Raudenbush, 1987) (Steele, 2008) Briefly, multilevel growth-curve models estimate population-averaged trajectories that quantify how a trait changes over time. Descriptive statistics and previous research indicated that depressive symptoms followed a non-linear pattern, (Kwong et al., 2019) so we used a quartic polynomial model to account for this non-linearity. The trajectory models were therefore comprised of an intercept term and four age terms (age, age^2^, age^3^ and age^4^).

To examine trajectories stratified by maternal and paternal maladaptive personality traits, we first created binary dummy variables to indicate the presence of maternal and paternal maladaptive personality traits and interacted these with all fixed effect terms (intercept and four age terms). Interpreting raw coefficients corresponding to the transformation of age terms is extremely difficult, therefore to interpret group differences we compared predicted depressive symptoms scores for offspring exposed and unexposed to maladaptive parental personality traits at ages 10, 14, 18 and 22. Separate models were used to estimate the effect of maternal and paternal personality. Full details are provided in Supporting information.

#### Modification of parental maladaptive personality traits association with offspring depression by SEP

To assess interactions, we created dummy variables for each level of the interaction between parental personality and each SEP indicator (exposed + high SEP, exposed + low SEP, not exposed + high SEP, not exposed + low SEP). These dummy variables were then interacted with the fixed effect terms as above. Effect modification was tested by comparing the difference in depressive symptoms for offspring exposed and unexposed to maladaptive parental personality within strata of high and low SEP at four key ages (10, 14, 18 and 22).

Multi-level modelling was conducted using Stata 15 (StataCorp, College Station, TX, USA) using the user-written runmlwin command (Leckie & Charlton, 2012), which calls the standalone multilevel modelling package MLwiN v3.02 (http://www.cmm.bristol.ac.uk/MLwiN). All figures were plotted using the ggplot2 package in R version 3.51.

#### Missing data

Available data for depressive symptoms decreased over time: 47% of the sample had non-missing data at age 11 which decreased to 25% by age 9 (Table S2). We included individuals with at least one measurement of depressive symptoms and complete data for maladaptive personality, which consisted of 7046 mother-child pairs and 3054 father-child pairs. Missing data was handled using full information maximum likelihood estimation (FIML).

## Results

Table 1 shows sample characteristics of mothers and fathers with low and high levels of maladaptive personality traits. Fifteen percent of mothers and 11 per cent of fathers were categorised as having high levels of maladaptive traits. Similar patterns were observed for both: mothers and father with high trait levels were more likely to have lower education, to have experienced financial problems, to have smoked in/around the time of their partner’s pregnancy and to have experienced intimate partner violence. They also both showed higher levels of depression in the post-natal period. We observed a low correlation between maladaptive maternal and paternal personality (r = 0.10).

**Table 1:** Sample characteristics of mothers and fathers

|  | Low maternal maladaptive traits |  | High maternal maladaptive traits |  |  |
| --- | --- | --- | --- | --- | --- |
|  | Mean (sd) | N (%) | Mean (sd) | N (%) | Odds ratio (95 % CI) / Mean |

|  |  |  |  |  | difference |
| --- | --- | --- | --- | --- | --- |
| **Mothers** | **N=6002 (85%)** |  | **N=1044 (15%)** |  |  |
| High education |  | 2584 (0.46) |  | 356 (0.37) | 0.71 (0.62 - 0.82) |
| Material hardship |  | 778 (0.14) |  | 364 (0.07) | 1.11 (1.09 - 1.13) |
| Age | 29.35 (4.42) |  | 28.37 (4.92) |  | -0.98 |
| Drinking in pregnancy |  | 3180 (0.56) |  | 564 (0.59) | 1.13 (0.98 - 1.30) |
| Smoking in pregnancy |  | 874 (0.15) |  | 279 (0.29) | 2.26 (1.93 - 2.64) |
| Depression in post-natal period |  | 391 (0.07) |  | 175 (0.19) | 3.04 (2.51 - 3.70) |
| Experienced partner violence |  | 209 (0.04) |  | 91 (0.10) | 2.81 (2.18 - 3.64) |
| **Fathers** | **n=2726 (89%)** |  | **n=328 (11%)** |  |  |
| High education |  | 1560 (0.63) |  | 139 (0.51) | 0.60 (0.47 - 0.77) |
| Material hardship |  | 391 (0.17) |  | 165 (0.07) | 1.15 (1.11 - 1.19) |
| Age | 31.04 (6.63) |  | 30.01 (7) |  | -1.03 |
| Smoking in pregnancy |  | 581 (0.25) |  | 107 (0.41) | 2.09 (1.61 - 2.72) |
| Depression in post-natal period |  | 58 (0.02) |  | 26 (0.1) | 4.43 (2.74 - 7.18) |
| Experienced partner violence |  | 70 (0.03) |  | 22 (0.08) | 3.04 (1.85 - 4.99) |

Note: Descriptive data shown for participants with complete data for exposure (parental personality) and at least one depression outcome (SMFQ at one time point). Odds ratios represent probability of given characteristic for high vs low maladaptive personality group. Mean differences represent mean score for high maladaptive personality group minus mean score for low maladaptive personality group.

### Aim 1: To describe trajectories of depressive symptoms of offspring with parents with and without high levels of maladaptive personality traits

Figure 1 shows the trajectories of depressive symptoms for offspring of mothers and fathers with and without high levels of maladaptive personality traits. Full model results are presented in Tables S3 – S6. The shape of the trajectories for offspring exposed and unexposed to parental maladaptive personality traits and in relation to each parent were similar, with increases in depressive symptoms to age 17–18 years followed by a slower decline to age 22–23. At both ends of the trajectories there were short ‘upturns’ but these may be influenced by sparse data at these ages. For mothers, there was evidence that the presence of maladaptive personality traits was associated with higher levels of offspring depressive symptoms across the trajectory from age 11 to 24 (predicted SMFQ difference in mean comparing those whose mothers had maladaptive personality traits to those who did not: age 10 = 0.66, 95% CIs 0.25, 1.28; age 14 = 0.31, 95% CIs 0.02, 0.61; age 18 = 0.96, 95% CIs 0.56, 1.37; age 22 = 1.00, 95% CIs 0.51, 1.50).

**Figure 1:**
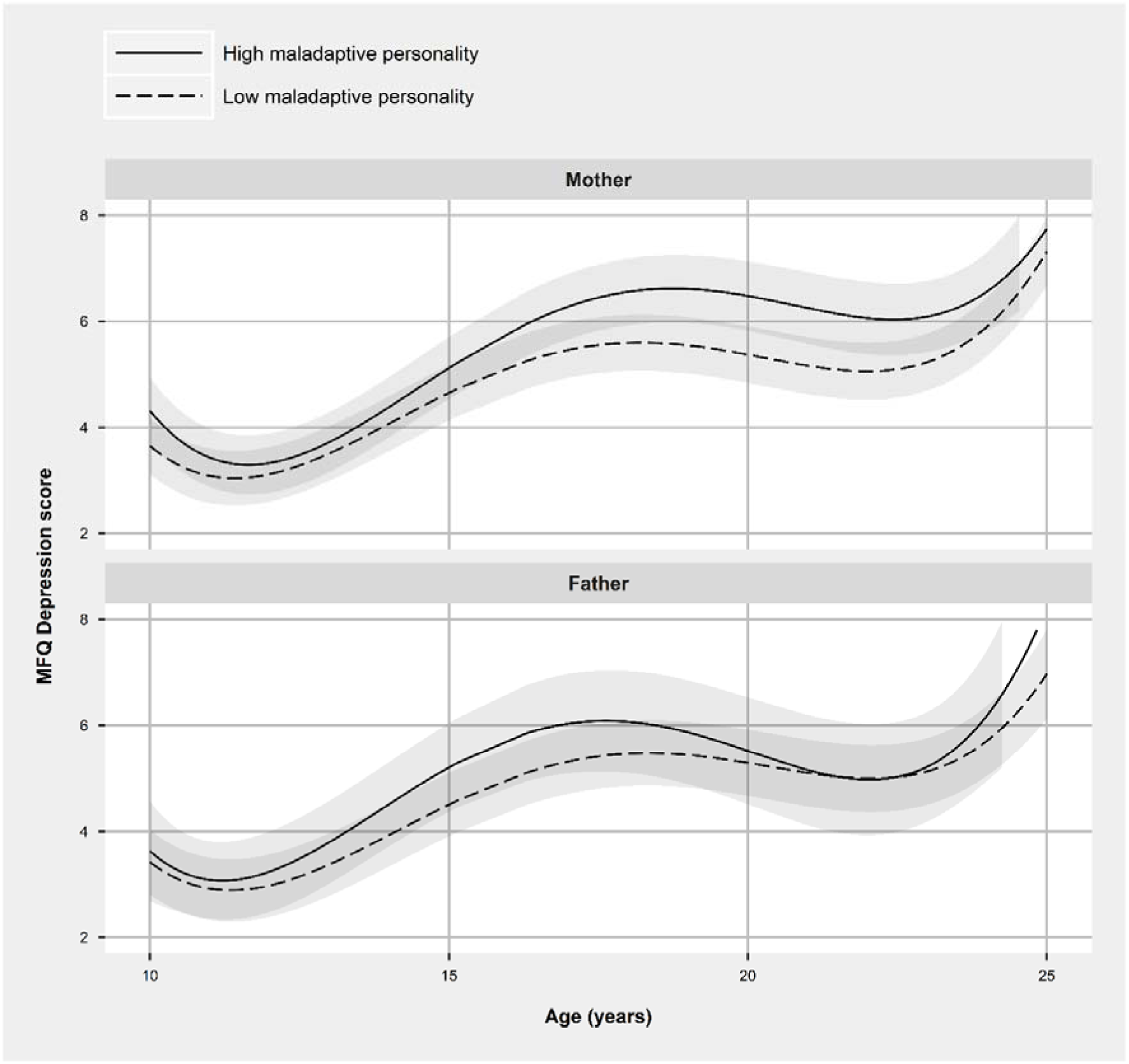
Trajectories of depressive symptoms for offspring by presence or absence of high maladaptive personality traits in mothers (N = 6232) and fathers (N = 2098)

Associations of paternal maladaptive personality traits with trajectories of offspring depressive symptoms were weaker and close to null by age 22 (predicted difference age 10 = 0.21, 95% CIs –0.58, 0.99; age 14 = 0.58, 95% CIs-0.01, 1.17; age 18 = 0.60, 95% CIs –0.21, 1.41; age 22 = 0.02, 95% CIs –0.94, 0.90); however it should be noted the paternal sample size is considerably smaller than that for maternal analyses and the confidence intervals for the two trajectories overlapped.

### Aim 2: To examine whether associations of parental personality on offspring depression are moderated by SEP

To test whether the relationship between maternal personality and offspring depressive symptoms are moderated by SEP, two separate analyses were completed for maternal education and material hardship. The shape of the trajectories across strata were similar with increases in depressive symptoms for all groups to age 17–18 followed by a decline to age 22–23. For both indicators of SEP, there was evidence that at age 18 the highest levels of depressive symptoms were in the high maladaptive personality x low SEP group; however at ages 10 and 24 scores were very similar for all groups (Figure 2 & Tables S7-S10).

**Figure 2:**
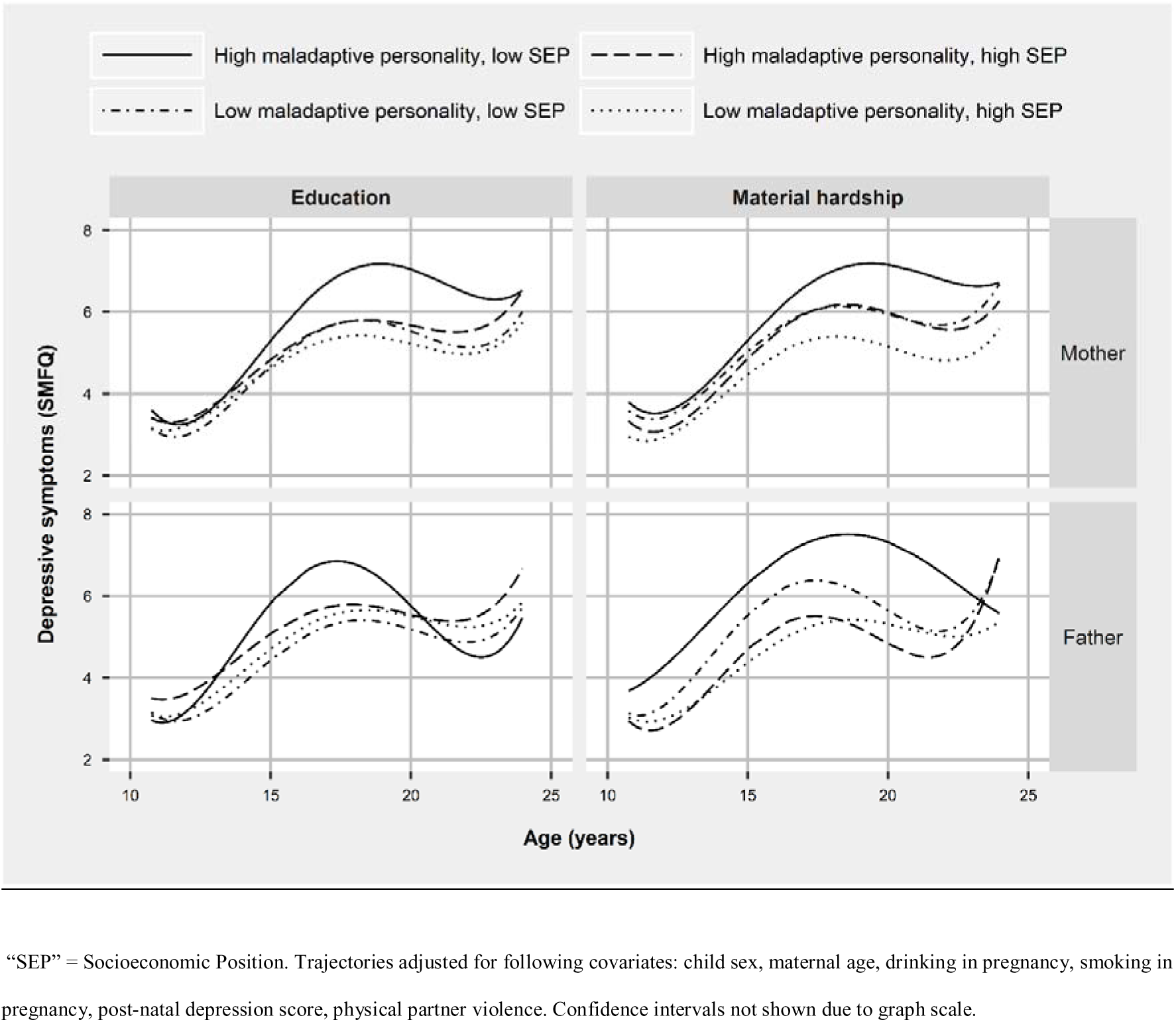
Trajectories of depressive symptoms stratified by socioeconomic position

Effect modification was tested by comparing the predicted difference in depressive symptoms for offspring exposed and unexposed to maladaptive parental personality within strata of high and low SEP at four key ages (10, 14, 18 and 22). We found minimal statistical evidence for an interaction between SEP and either maternal or paternal personality, with only two out of sixteen tests reaching the threshold of statistical significance (Table 2). For example, at age 10 the predicted difference in mean SMFQ comparing those exposed to maladaptive maternal personality to those who were not was 0.42 (−0.23, 1.07) in those whose mothers had higher education and 0.67 (0.13, 1.20) in those whose mothers had lower education, P-interaction = 0.56.

**Table 2:**
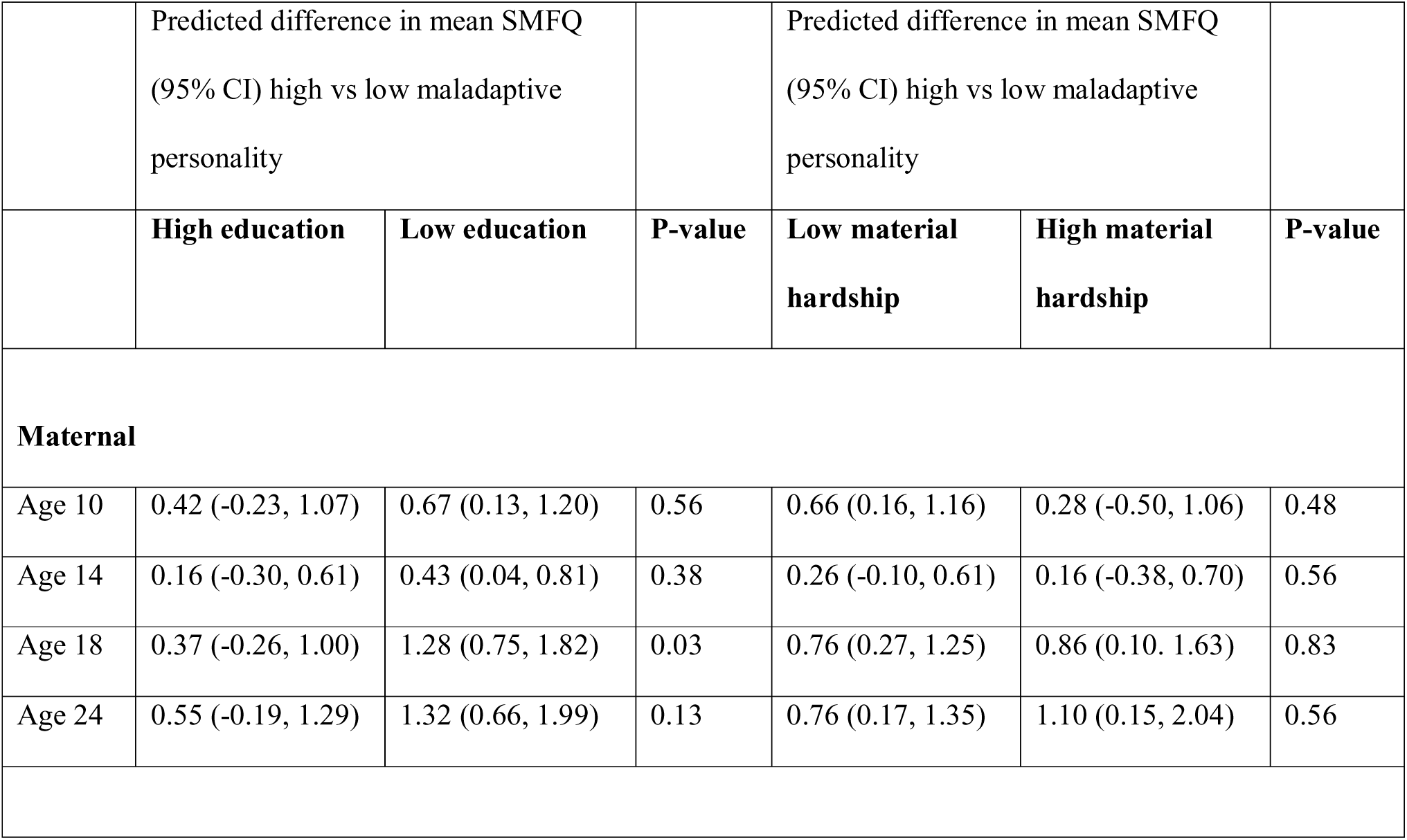
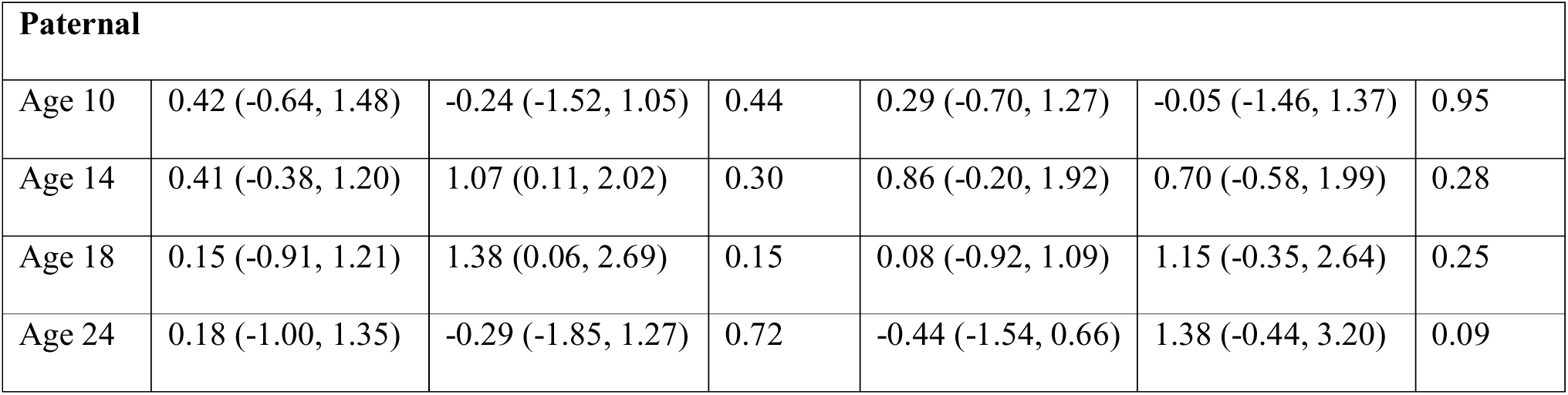
Difference in depressive symptoms comparing offspring exposed and unexposed to maladaptive parental adaptation within strata of high and low SEP

|  |  |  |  |  |  |  |
| --- | --- | --- | --- | --- | --- | --- |
|  | Predicted difference in mean SMFQ<br>(95% CI) high vs low maladaptive<br>personality |  |  | Predicted difference in mean SMFQ<br>(95% CI) high vs low maladaptive<br>personality |  |  |
|  | **High education** | **Low education** | **P-value** | **Low material**<br>**hardship** | **High material**<br>**hardship** | **P-value** |
| **Maternal** |  |  |  |  |  |  |
| Age 10 | 0.42 (-0.23, 1.07) | 0.67 (0.13, 1.20) | 0.56 | 0.66 (0.16, 1.16) | 0.28 (-0.50, 1.06) | 0.48 |
| Age 14 | 0.16 (-0.30, 0.61) | 0.43 (0.04, 0.81) | 0.38 | 0.26 (-0.10, 0.61) | 0.16 (-0.38, 0.70) | 0.56 |
| Age 18 | 0.37 (-0.26, 1.00) | 1.28 (0.75, 1.82) | 0.03 | 0.76 (0.27, 1.25) | 0.86 (0.10, 1.63) | 0.83 |
| Age 24 | 0.55 (-0.19, 1.29) | 1.32 (0.66, 1.99) | 0.13 | 0.76 (0.17, 1.35) | 1.10 (0.15, 2.04) | 0.56 |

| **Paternal** |  |  |  |  |  |  |
| --- | --- | --- | --- | --- | --- | --- |
| Age 10 | 0.42 (-0.64, 1.48) | -0.24 (-1.52, 1.05) | 0.44 | 0.29 (-0.70, 1.27) | -0.05 (-1.46, 1.37) | 0.95 |
| Age 14 | 0.41 (-0.38, 1.20) | 1.07 (0.11, 2.02) | 0.30 | 0.86 (-0.20, 1.92) | 0.70 (-0.58, 1.99) | 0.28 |
| Age 18 | 0.15 (-0.91, 1.21) | 1.38 (0.06, 2.69) | 0.15 | 0.08 (-0.92, 1.09) | 1.15 (-0.35, 2.64) | 0.25 |
| Age 24 | 0.18 (-1.00, 1.35) | -0.29 (-1.85, 1.27) | 0.72 | -0.44 (-1.54, 0.66) | 1.38 (-0.44, 3.20) | 0.09 |

## Discussion

Knowledge about the influence of parental personality on offspring mental health is limited. We found that the presence of maladaptive personality traits among mothers was associated with higher levels of offspring depressive symptoms across the trajectory from age 11 to 24. Yet, the shape of the trajectories for offspring exposed and unexposed to parental maladaptive personality traits were similar, with increases in depressive symptoms to age 17–18 years followed by a slower decline to age 22–23. The associations of paternal maladaptive personality traits with trajectories of offspring depressive symptoms were weaker and close to null by age 22, though the paternal sample size was considerably smaller than that for the maternal analyses.

At age 18 we found that the highest levels of depressive symptoms were for offspring of mothers with both high levels of maladaptive traits and low SEP. However, there was no statistical evidence for an interaction suggesting that these are risk factors are independent from one another and the greater risk observed in offspring exposed to both maladaptive personality and low education occurs due to the additive accumulation of two factors.

### Trajectories of depressive symptoms

The offspring of mothers with high levels of maladaptive personality traits had trajectories that were higher across adolescence and young adulthood, with the differences increasing from approximately age 15 and the greatest difference occurring around the age of 22. The magnitude of these effects was small, equating to a difference in SMFQ scores of approximately 1 at age 18 (SMFQ range = 0 – 26). This extends our previous findings that clinical levels of depression at 18 are more frequent amongst those exposed to maladaptive maternal personality traits. (Pearson at al. 2018) It also suggests that adolescence is a period of high risk for this group of offspring, as symptoms do not necessarily decrease to population average levels.

In contrast we found a smaller effect of father’s personality on the level of offspring depressive symptoms. However, it is important to note that the sample size for paternal trajectory analysis was considerably smaller, and that for all estimates the confidence intervals for associations between maternal and paternal personality and offspring depression overlapped.

There is evidence that depression and maladaptive personality traits share common genetic correlates, (Witt et al., 2017) therefore part of the association between maternal maladaptive personality and offspring depression will be accounted for by shared genotype. It is also likely that some of the association between parental personality and offspring depression is mediated through environmental factors. This is supported by our finding of a weaker association of paternal personality with offspring depression, as if the effect was entirely genetic we would expect to see equivalent associations with maternal and paternal personality. Although not tested in this study, a plausible mediating factor for the association is through the parent-child relationship. This would be consistent with evidence of less sensitive parenting in mothers with higher levels of maladaptive personality traits (Eyden et al., 2016) and the prospective association between parenting style and offspring depression. (Stein et al., 2014)

It also possible that there are evocative gene-environment correlations: given that personality traits are partly heritable (Distel et al., 2011) offspring of parents with more difficult and maladaptive personality traits are also more likely to have such traits, which in turn could evoke a harsher parenting style or conflicts. For example, having an impulsive and aggressive child may evoke more hostile parental responses which in turn might lead to unhappiness in the child. Future studies including a genetic component could test this hypothesis.

If the finding of a stronger effect of maternal vs paternal personality on offspring depression is replicated, it could be explained by cultural differences in maternal and paternal parenting and/or quantity of time spent with mothers due to breastfeeding, higher frequency of non-working or part time working mothers vs fathers in the UK at the time of the study. However the notion of differential effects of mothers and father parenting on the child is increasingly questioned given the limited empirical evidence that the parenting of mothers and fathers is qualitatively different. (Cabrera, Fagan, Wight, & Schadler, 2011) (Fagan, Day, Lamb, & Cabrera, 2014) Future studies should therefore attempt to replicate these findings of a weaker effect of paternal vs maternal personality on offspring depression, and test whether any association between parental personality and offspring depression is mediated by parenting, relational factors or other indirect effects of traits such as impulsivity (e.g. a more chaotic home environment).

### Effect modification by socioeconomic position

Whilst we consistently found that the highest levels of offspring depressive symptoms were in the high maladaptive traits, low SEP groups, there was minimal statistical evidence of effect modification. This is consistent with a recent large study which reported no evidence of an interaction between maternal depression and SEP on offspring depression, (Mikkonen et al., 2016) but not with a previous ALSPAC study which did report an interaction between maternal post-natal depression and SEP on offspring depression. (Pearson et al., 2013). These inconsistencies may be a result of the large sample required to detect interaction effects, but also suggest that if there is an interaction effect it is likely a very small one. (Kendler & Gardner, 2010).

The absence of effect modification is potentially an important clinical finding. It suggests that whilst both SEP and maladaptive parental personality are risk factors for offspring depression, the effect of parental personality traits such as high impulsivity or suspicion are the same across low and high SEP families. This finding was consistent using two separate indicators representing separate (though likely overlapping) aspects of SEP. (Galobardes et al., 2006) This implies that there is unlikely to be a clinically unique group of ‘at risk’ parents with low SEP and high levels of maladaptive traits which require targeting. Instead interventions are likely to be required on different levels, e.g. policy and economic interventions to reduce inequality vs family and individual level interventions for parents with maladaptive personality traits.

### Summary and implications

This study has provided converging evidence that the offspring of mothers with high levels of maladaptive personality traits show a greater increase of depressive symptoms through adolescence, although the magnitude of the effect modest. A similar trend was observed for fathers but to a lesser degree, notwithstanding the considerably smaller paternal sample for the trajectory analysis. Thus, this describes two vulnerable groups: (i) mothers with high levels of maladaptive traits, many of whom have experienced interpersonal violence and mental health problems themselves, (Moran et al., 2016) and (ii) their offspring who are at increased risk of depression throughout adolescence. If replicated these results may point to the potential need for early interventions for these families which take account of the interpersonal style and personality characteristics of the parents.

### Strengths and limitations

This was a large prospective cohort study with extensive repeated data on depressive symptoms. Extensive data on potential confounders was available and included in the analysis. However, it is still likely that residual confounding was present as traits such as personality and SEP indicators will be associated with a wide range of other individual and social factors. A key limitation of this study was the smaller sample size for the paternal trajectories compared to the maternal analysis, which limits the opportunity for comparison of associations between parents. A further limitation of the findings is the high attrition rate within ALSPAC, which is associated with sex and socioeconomic factors. (L D Howe, Galobardes, Tilling, & Lawlor, 2011) However, our use of FIML would have partially mitigated this bias. Whilst our use of polynomial models allowed the estimation of natural growth curves, interpretation of age coefficients was difficult and future studies modelling mental health trajectories could consider spline approaches. (L. D. Howe et al., 2016) Finally, notwithstanding the reasonable sample size, cell numbers for some of the interaction levels were still small and standard errors high.

### Future directions

These findings need to be replicated in an independent larger sample, to confirm whether there are differences in associations between mothers and fathers with maladaptive traits and offspring depressive symptoms. To understand the potential mechanisms of transmission potential mediating pathways of the association should be tested (e.g. later parent-child relationships or parental depression), and potential genetic effects, including evocative, also explored.

## Data Availability

Data is not publicly available, however any researchers are able to apply for use of ALSPAC data.

## Acknowledgements

We are extremely grateful to all the families who took part in this study, the midwives for their help in recruiting them, and the whole ALSPAC team, which includes interviewers, computer and laboratory technicians, clerical workers, research scientists, volunteers, managers, receptionists and nurses.

<u>Correspondence to</u>: Tim Cadman, Integrative Epidemiology Unit (IEU), University of Bristol, Oakfield House, Oakfield Grove, Bristol, BS8 2BN.

## Abbreviations

SMFQ: “Short mood and feelings questionnaire”

